# Causal Relationship between Rheumatoid Arthritis and Atherosclerosis Risk: A Mendelian Randomized Study

**DOI:** 10.1101/2024.04.14.24305792

**Authors:** Qi-Pei Liu, Hong-Cheng Du, Ping-Jin Xie, Lei-Xiao Zhang, Hao-Ze Gao, Jie-Hua Luo, Sheng-Ting Chai

## Abstract

**Objectives:** Previous observational studies have revealed an association between rheumatoid arthritis (RA) and atherosclerosis (AS). However, observational studies are prone to bias due to potential confounding factors and reverse causality. Therefore, this study investigates the causal relationship between RA and AS risk using Mendelian randomization (MR).

**Methods:** Genetic data related to RA and AS were sourced from the Integrative Epidemiology Unit Open-Genome-Wide Association Studies database and the seventh edition of the FinnGen Biobank gene database, respectively. The initial analysis employed the inverse variance weighted (IVW) method for the MR analysis, with other methods utilized as supplementary methods. To ensure the robustness and reliability of the conclusions, various sensitivity analyses were conducted.

**Results:** The MR results indicate a positive causal relationship between genetically determined RA and the risk of coronary atherosclerosis(OR = 1.026, 95% CI: 1.004, 1.049, p = 0.179), peripheral atherosclerosis(OR = 1.072, 95% CI: 1.035, 1.111, p < 0.001), and atherosclerosis excluding coronary atherosclerosis, cerebral atherosclerosis, and PAD (ASE) (OR = 1.046, 95% CI: 1.015, 1.079, p = 0.004), while no significant causal relationship with cerebral AS was observed. The MR-Egger regression did not reveal any significant horizontal pleiotropy.

**Conclusion:** This study elucidates the causal relationship between RA and various AS risks, highlighting the importance of actively detecting and intervening in RA for AS risk.

## Introduction

Rheumatoid arthritis (RA) is an autoimmune disease characterized by persistent synovitis, cartilage, and bone degradation. Its main clinical manifestations include joint pain, swelling, stiffness, deformity, and sleep disturbances^1,2^. The prevalence of RA in most countries worldwide typically ranges from 0.25% to 3.4%^3^. The male-to-female ratio is 1:3, and the age of onset is primarily concentrated between 50 and 60 years^4,5^. From 1990 to 2019, the global age-standardized prevalence rate (ASPR) of RA significantly increased, and the ASPR in Europe and the Americas is generally greater than that in Africa and Asia^6^.

Atherosclerosis (AS) is a chronic inflammatory disease characterized by the deposition of lipids beneath the arterial intima. Mechanisms such as inflammatory responses, endothelial dysfunction, abnormal immune responses, and disrupted lipid metabolism can lead to the development of atherosclerosis or atheromatous plaques^7^. Under the influence of metalloproteinases, inflammatory cell infiltration, and other factors, the plaque is prone to rupture, thereby triggering acute cardiovascular diseases such as ischemic stroke and myocardial infarction^8^. Depending on the affected arterial location, AS can be classified into coronary atherosclerosis, peripheral artery atherosclerosis, cerebrovascular artery atherosclerosis, renal artery atherosclerosis, mesenteric artery atherosclerosis, pulmonary artery atherosclerosis, and so on.

Observational studies investigating the relationship between RA and AS have yielded conflicting conclusions and are susceptible to bias due to unknown confounding factors and reverse causality. Mendelian randomization (MR) is an epidemiological method for causal analysis that utilizes genetic variation as instrumental variables (IVs)^9^. Since environmental factors and the disease process do not influence genetic variations, MR can mitigate reverse causality and reduce the influence of confounding factors, thereby strengthening the relationship between exposure and outcome^10^. Although previous MR studies have provided evidence showing that genetically predicted RA increases the risk of coronary atherosclerosis^11^, there is a lack of research on other more widespread atherosclerotic conditions such as peripheral artery atherosclerosis, cerebral artery atherosclerosis, and so on. Furthermore, to our knowledge, there is currently no MR study investigating the causal impact of genetic susceptibility to RA on various forms of atherosclerosis. Therefore, in our study, univariable Mendelian randomization (UVMR) analysis was implemented to explore the correlation between RA and various types of AS, including coronary AS, cerebral AS, peripheral AS, and atherosclerosis excluding coronary atherosclerosis, cerebral atherosclerosis, and PAD (peripheral arterial disease) (ASE), to further enrich and supplement the theoretical basis.

## Methods and Materials

### Study design

To investigate the relationship between RA and AS risk, we conducted UVMR analysis and reverse UVMR analysis using single nucleotide polymorphisms (SNPs) as IVs^12^. The flowchart of the study can be found in Supplementary Figure S1. Three crucial assumptions must be met to ensure the accuracy of the study^13^. First, the SNPs included in the study were significantly associated with RA. Second, the SNPs were independent of confounding factors related to both RA and AS. Third, all SNPs act on AS only through RA (Supplementary Figure S1).

### Data sources

The summary-level statistical data of genetic variations related to RA included in our study were obtained from the publicly available Integrative Epidemiology Unit (IEU) Open-Genome-Wide Association Studies (OpenGWAS) database^14^, which includes a total of 58,284 samples involving both males and females and 13,108,512 SNPs ^15^(Supplementary Table S1). The summary statistical data for genetically determined coronary AS, cerebral AS, peripheral AS, and ASE are derived from the seventh edition of the FinnGen Biobank gene database^16,17^ (Supplementary Table S1). To mitigate population stratification bias, all SNPs included in this study were derived from studies conducted on individuals of European ancestry.

All data included in the study are accessible from the publicly available IEU OpenGWAS database and the FinnGen Biobank genetic database. Therefore, ethical approval and informed consent were not required for this study.

### IVs selection

Firstly, we selected SNPs significantly associated with the entire genome of RA exposure (range of p < 5*10^^-8^). For reverse UVMR, a threshold of p < 1*10^^-5^ was set to include more SNPs related to AS. Secondly, to ensure data validity, we applied a linkage disequilibrium (LD) clustering process (r^2^ < 0.001, clustering distance cutoff = 10,000 kb) to estimate the LD between the selected SNPs^18^. Finally, to test whether the selected genetic variations as IVs have a weak association with the exposure, the F-statistic was calculated. To avoid weak instrument bias, IVs with F<10 were excluded.

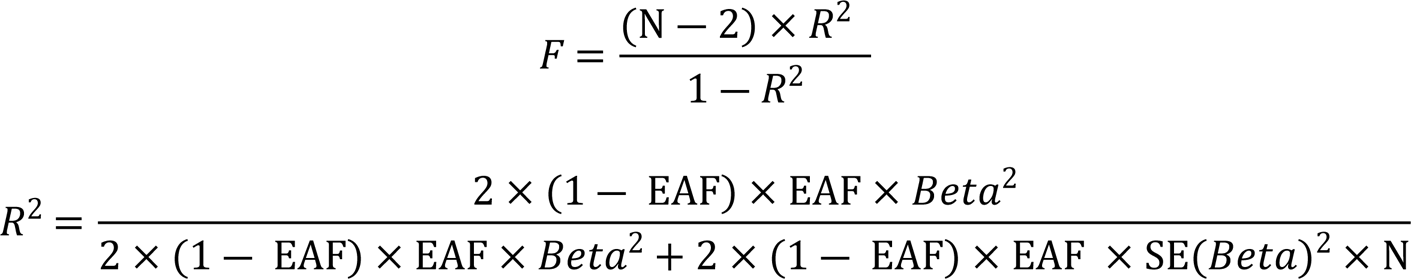

Beta represents the estimated genetic effect, EAF denotes the effect allele frequency, N indicates the sample size, and SE(Beta) denotes the standard error of the genetic effect^19^.

### Statistical analysis

All the data were analyzed using the TwoSampleMR package in RStudio (version 4.2.1)^20^.

### MR analysis

The inverse variance weighted (IVW) method is typically used as the most reliable MR analysis method to explore causal effects^21^. Since the IVW method assumes that all SNPs are valid instruments, weighted median (WM) and MR-Egger were also included as supplementary methods for MR analysis to assess the reliability of the results. MR-Egger is susceptible to the influence of pleiotropic genetic variations, allowing for causal effect estimates of lower precision under weaker assumptions for the included genetic variations^22^. The WM method requires more than 50% effectiveness among the included IVs and estimates causal effects based on the weights of different genetic variations, making it more accurate than estimates based on the MR-Egger regression method^23^. The results based on the IVW method were statistically significant and consistent in direction with the results based on the WM method or MR-Egger regression, indicating significant causal effects in our study.

### Sensitivity analysis

First, the presence of significant horizontal pleiotropy in IVs and other potential confounding factors were assessed through MR-Egger regression^24^. Second, the IVW and MR-Egger regression were conducted to detect the presence of heterogeneity and quantify it using the Cochrane Q statistic^25^. A random-effects model was used for IVW when significant heterogeneity was present. Otherwise, a fixed-effects model was applied for IVW. Third, the symmetry of the funnel plot was analyzed to assess horizontal pleiotropy, indicating a lower likelihood of potential bias risk affecting the study conclusions when the plot is symmetrical^26^. Finally, leave-one-out analysis was conducted by systematically removing one IV at a time to assess whether the remaining IVs significantly affected the causal effect, thus evaluating whether any IV biased the conclusion.

## Results

### Results of the Selection of IVs

For detailed information about the SNPs used in the genetic IVs, as well as their associations with RA, coronary AS, cerebral AS, peripheral AS, and ASE, please refer to Supplementary Data S1.

### Effects of RA on AS risk

Based on IVW, MR-Egger regression, and WM analyses, no significant causal relationship was found between genetic susceptibility to RA and cerebral AS risk (OR = 0.920, 95% CI: 0.800, 1.057, p = 0.237) (Supplementary Figure S2). MR based on IVW estimates showed a strong correlation between genetically determined RA and coronary AS (OR = 1.026, 95% CI: 1.004, 1.049, p = 0.179), peripheral AS (OR = 1.072, 95% CI: 1.035, 1.111, p < 0.001) and ASE (OR = 1.046, 95% CI: 1.015, 1.079, p = 0.004) (Supplementary Figure S2). MR-Egger regression and WM also showed similar trends (Supplementary Figure S2). Plotting the SNP effects of selected RA (X-axis) against the SNP effects of coronary AS, cerebral AS, peripheral AS, and ASE (Y-axis) indicated a significant positive causal effect of RA overall and coronary AS, peripheral AS, and ASE SNP effects (Supplementary Figure S3).

### Results of the sensitivity analysis

In the MR analysis involving genetically determined RA and coronary AS, peripheral AS, and ASE, significant heterogeneity was observed (Supplementary Table S2). All MR-Egger analyses of the risk of RA and AS in the forest plots did not intersect with zero (Supplementary Figure S4). MR-Egger regression was not statistically significant (Supplementary Table S2). Additionally, by examining the funnel plot (Supplementary Figure S5), it can be observed that the distribution of all included SNPs around the IVW estimated values is generally symmetrical. Furthermore, the “leave-one-out” analysis did not identify any IVs as outliers (Supplementary Figure S6). All SNPs included in our study had an F-statistic > 10 (Supplementary Data S1).

### Results of reverse MR analysis

Based on the reverse UVMR analysis using the IVW method, MR-Egger regression, and WM method, no significant causal effect was detected between genetically determined coronary AS, cerebral AS, peripheral AS, ASE, and RA risk (Supplementary Table S3). MR-Egger regression did not reveal any significant horizontal pleiotropy (Supplementary Table S4). All SNPs included in the study were computed with an F-statistic > 10 (Supplementary Data S1).

## Discussion

Through this study, it was found that the genetic susceptibility to rheumatoid arthritis is positively correlated with the risk of coronary AS, peripheral AS, and ASE, with no significant association observed with cerebral AS risk. Moreover, no significant genetic correlation was detected in the reverse MR study between coronary AS, cerebral AS, peripheral AS, ASE, and RA risk. Furthermore, the reliability and stability of the conclusions were validated through various sensitivity analyses.

Previous studies have shown that the onset of atherosclerosis precedes that of RA^27^, and there is an increasing belief that atherosclerosis is becoming a primary cause of rheumatoid arthritis^28^. In contrast, several cross-sectional studies have found that the incidence of coronary atherosclerosis, peripheral atherosclerosis, aortic valve sclerosis, and carotid atherosclerosis in RA patients is greater than that in the general population, sometimes even doubling^29–32^. Furthermore, a meta-analysis revealed that compared to the general population, RA patients exhibit greater carotid intima-media thickness (CIMT), along with a significantly increased incidence of carotid atherosclerosis (OR: 3.61; 95% CI: 2.65, 4.93; p < 0.0001)^29^. Our MR analysis results support that RA increases susceptibility to various forms of AS, including coronary atherosclerosis, peripheral atherosclerosis, and ASE risk, but not cerebral atherosclerosis. Therefore, primary prevention of atherosclerosis in RA patients holds significant importance.

RA is an autoimmune disease that can lead to disability, and the mechanism by which it causes the onset of AS remains incompletely understood. Research has revealed that inflammatory cytokines such as IL-17, IL-1β, and TNF-α are widely present in RA patients^33,34^. These cytokines not only participate in synovial development but also promote the activation of endothelial cells, leading to dysfunction and accelerating the progression of atherosclerosis through the modulation of inflammatory cascades^33,35^. Moreover, RA chronic inflammation also has procoagulant and prooxidant effects, increasing the expression of tissue factor adhesion molecules, reducing the synthesis of nitric oxide and thrombomodulin, and inducing nicotinamide adenine dinucleotide phosphate oxidase-mediated mechanisms^36,37^. This further leads to endothelial dysfunction and shifts the hemostatic balance of RA toward a prothrombotic state. In addition, in RA patients, abnormal activation of T and B cells occurs, and CD4+ and Th17 cells can lead to abnormal secretion of inflammatory factors such as IL-17 and IL-6, triggering abnormal inflammatory responses and causing endothelial cell damage and dyslipidemia, ultimately leading to atherosclerosis^38,39^. The shared epitope allele combinations *HLA-DRB1*04*04* and *HLA-DRB1*01*04* have been demonstrated to not only be risk factors for promoting RA but also to be associated with endothelial dysfunction and carotid atherosclerosis^40,41^. Furthermore, studies have proposed the “lipid paradox” in RA patients, wherein serum levels of high-density lipoprotein (HDL), low-density lipoprotein (LDL), and total cholesterol decrease due to excessive degradation^37,42^. Additionally, a proinflammatory form of HDL, known as piHDL, is produced, which promotes foam cell formation and LDL oxidation, thus accelerating the progression of atherosclerotic plaque enlargement^43,44^. In summary, RA may trigger and exacerbate AS from various aspects, including inflammation, lipid disorders, endothelial dysfunction, immune dysfunction, and genetic factors.

Endothelial function and CIMT are recognized surrogate markers for early atherosclerosis^45^, and there is evidence suggesting that assessing skin vascular function can serve as a noninvasive, useful surrogate marker for evaluating vascular risk in RA^46^. RA medications such as the disease-modifying antirheumatic drugs (DMARDs) methotrexate (MTX)^47,48^ and the TNF-α inhibitors infliximab (IFX)^49–52^, adalimumab^52^, and rituximab^53^ have been clinically proven to reduce proinflammatory mediators such as IL-6 in RA patients and to consistently improve endothelial function, exerting beneficial effects on the development of AS in RA patients and thereby delaying the onset and mortality of cardiovascular disease. Moreover, the angiotensin-converting enzyme inhibitor ramipril can significantly improve endothelial function in patients with RA, offering a potential new strategy for preventing cardiovascular events in RA patients^54^. Moreover, clinical studies have demonstrated that long-term lipid-lowering therapy with statin drugs (such as rosuvastatin and atorvastatin) can improve endothelial function in RA patients by alleviating arterial stiffness and reducing carotid plaque height, thereby reducing the risk of cardiovascular events (CVEs)^55,56^.

This study has several strengths. Firstly, our research employs population restrictions on individuals of European descent to reduce population structure bias. Secondly, this MR study comprehensively covered various types of atherosclerotic diseases and strictly adhered to the STROBE-MR guidelines. To ensure the robustness and reliability of the conclusions, both IVW and two supplementary methods (WM and MR-Egger) were employed for MR analysis, along with various sensitivity analyses to mitigate potential confounding biases as much as possible. Although some heterogeneity was observed in the analysis of RA with AS from the IEU OpenGWAS database and the FinnGen dataset, the random-effects model could correct the heterogeneity. Furthermore, the MR-Egger regression analysis did not detect any significant horizontal pleiotropy, suggesting that the observed heterogeneity is unlikely to introduce bias to the MR analysis.

When analyzing our study results, certain limitations need to be considered. Firstly, all populations included in the MR study were of European descent, so the generalizability of the study conclusions to all ethnicities awaits future research. Secondly, potential unidentified confounding or mediating factors between exposure and outcome variables may introduce unpredictable biases to our study results. Third, the study faces a limitation as it cannot perform a stratified analysis based on essential variables such as sex. Lastly, due to the characteristics of Mendelian randomization studies, the assessment of potential nonlinear associations between RA and AS diseases is limited.

### Conclusion

In summary, this MR study revealed a positive correlation between genetic predispositions to RA and coronary AS, peripheral AS, and ASE, with no significant causal relationship observed with cerebral AS. This information can aid clinicians in identifying RA patients at greater risk of AS diseases, facilitating early diagnosis and comprehensive treatment. Additionally, in future clinical treatments, the use of drugs with potential benefits for targeted RA therapy, such as anti-TNF-α agents(e.g., etanercept, adalimumab, infliximab), which have proven potential benefits in AS treatment.

## Data Availability

All relevant data are within the manuscript and its Supporting Information files.The data underlying the results presented in the study are available from the publicly available Integrative Epidemiology Unit Open-Genome-Wide Association Studies database (https://gwas.mrcieu.ac.uk/) and the FinnGen Biobank genetic database (https://www.finngen.fi/en).

## Acknowledgments

We sincerely thank the participants and researchers from the FinnGen study and the UK Biobank, as well as James J. Lee et al. This work is based on the generous sharing of data from the IEU OpenGWAS database, the FinnGen Biobank genetic database, and contributions from James J. Lee and colleagues.

## Author contribution

Guiding the research process: ST C. Conceptualization and methodology, Study design, Statistical analysis, and Data collection: QP L and HC D. QP L and HC D these authors contributed equally to this work and shared first authorship. Writing (original draft & review & editing) and Approval of final manuscript: all authors.

## Data availability statement

All data included in this study can be obtained from the publicly available IEU OpenGWAS database and FinnGen Biobank gene database.

## Supplementary materials for

**Supplementary Figure SI.**
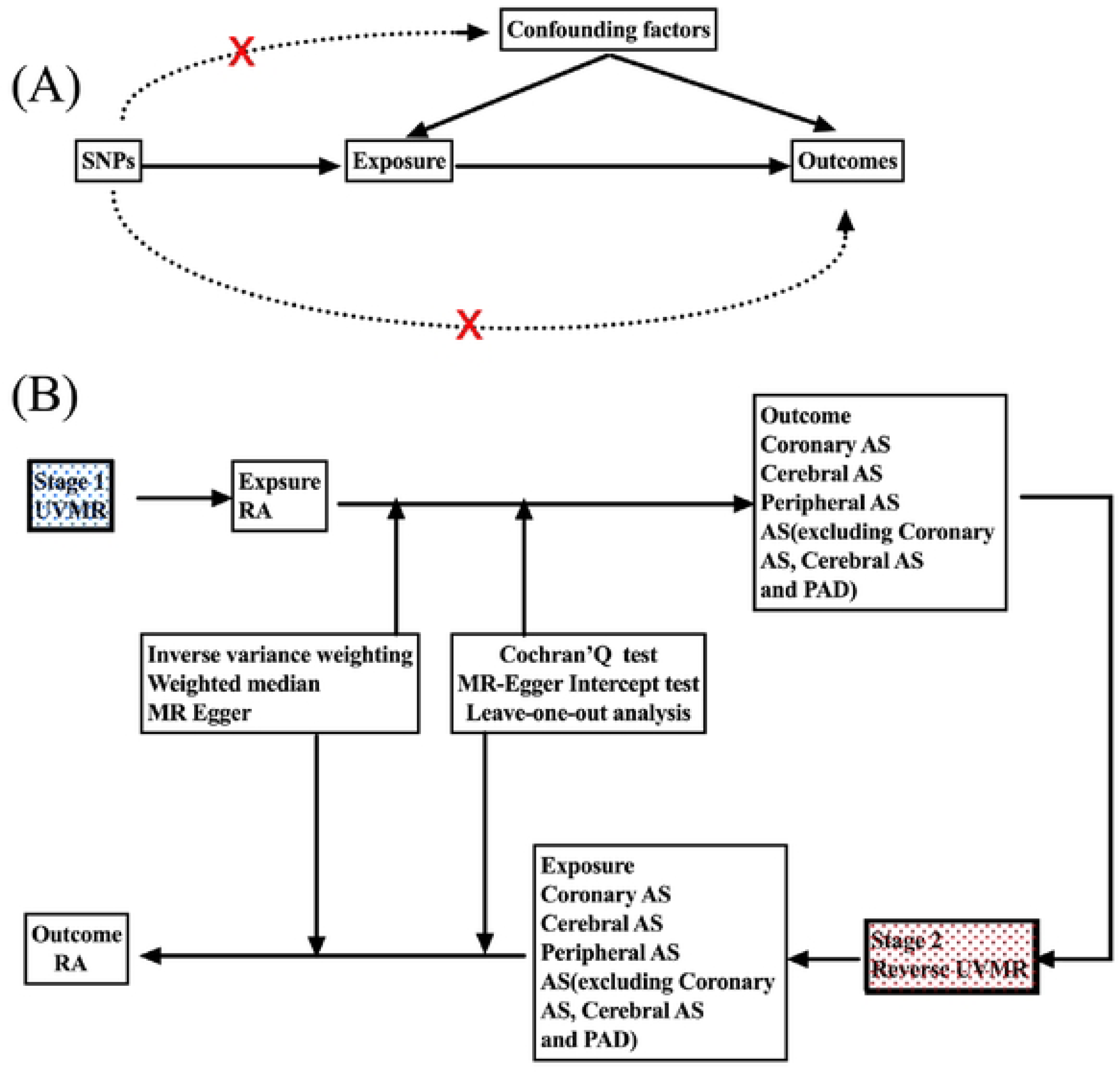
A working flow chart to explore the potential relationship between RA and AS risk. (A) The basic idea of MR Analysis; (B) the workflow of two-way UVMR Analysis. MR, Mendelian randomization; UVMR, Univariate Mendelian randomization; AS,atherosclerosis; RA, rheumatoid arthritis. SNPs, single nucleotide polymorphisms; PAD, peripheral arterial disease.

**Supplementary Figure S2.**
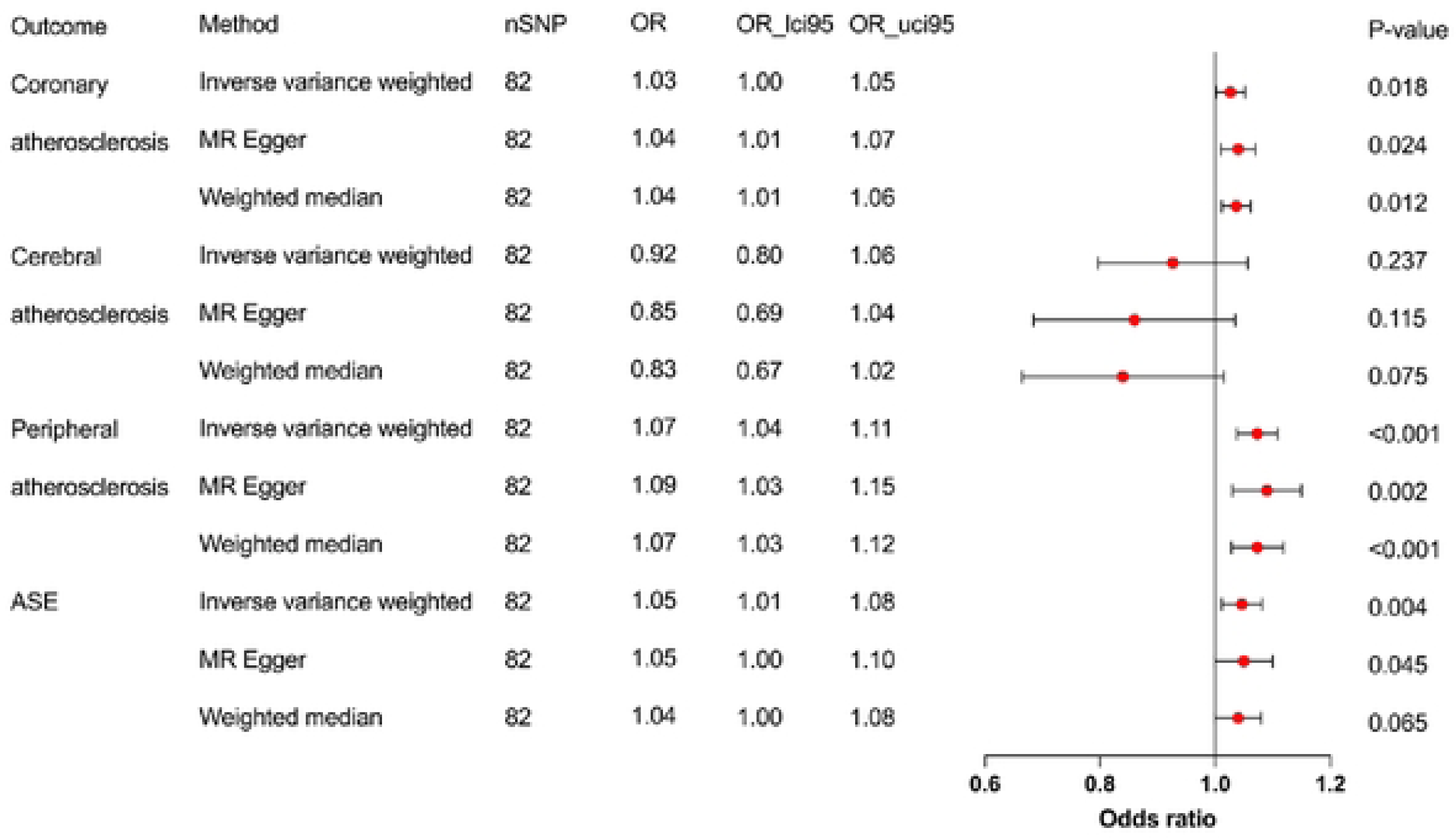
MR analysis of forest maps showing the effect of genetically determined RA on AS risk. nSNP, number of single nucleotide polymorphisms; OR, odds ratio; OR_lci95, The lower limit of the 95% confidence interval for the odds ratio; OR_uci95, The upper limit of the 95% confidence interval for the odds ratio; RA, rheumatoid arthritis; AS, atherosclerosis; ASE, atherosclerosis excluding coronary atherosclerosis, cerebral atherosclerosis, and PAD (peripheral arterial disease); MR, Mendelian randomization.

**Supplementary Figure S3.**
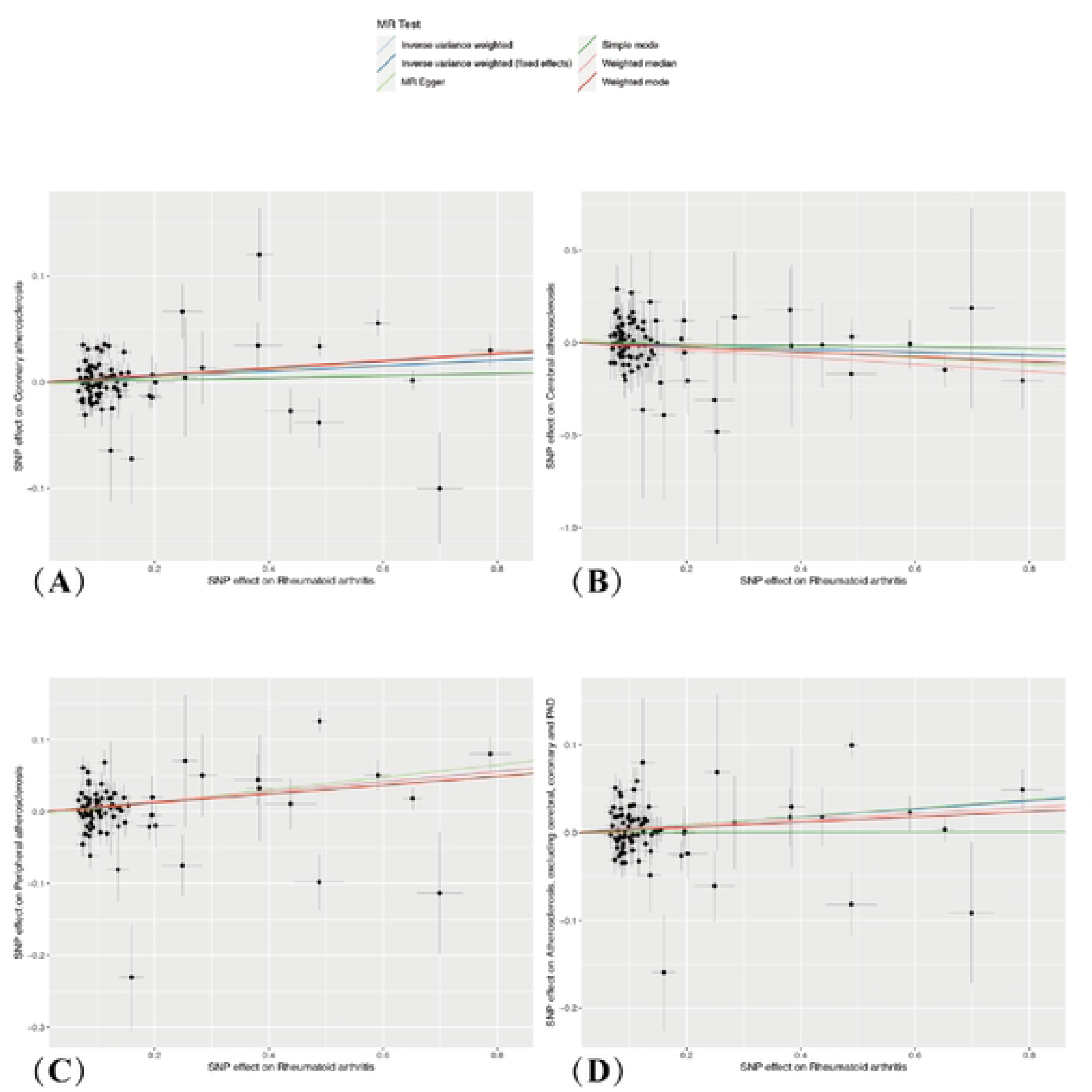
Scatterplot summary of genetically determined Causality between RA and AS under different MR methods. A: causal estimates for RA on coronary AS. B: causal estimates for RA on cerebral AS. C: causal estimates for RA on peripheral AS. D: causal estimates for RA on ASE. The slope of each line corresponds to the causal estimates for each method. The impact of each SNPs on the exposure is depicted using a point and a horizontal line, while its effect on the outcome is represented by a point and a vertical line. RA, rheumatoid arthritis; AS, atherosclerosis; ASE, atherosclerosis excluding coronary atherosclerosis, cerebral atherosclerosis, and PAD (peripheral arterial disease) SNPs, single nucleotide polymorphisms, MR, Mendelian randomization.

**Supplementary Figure S4.**
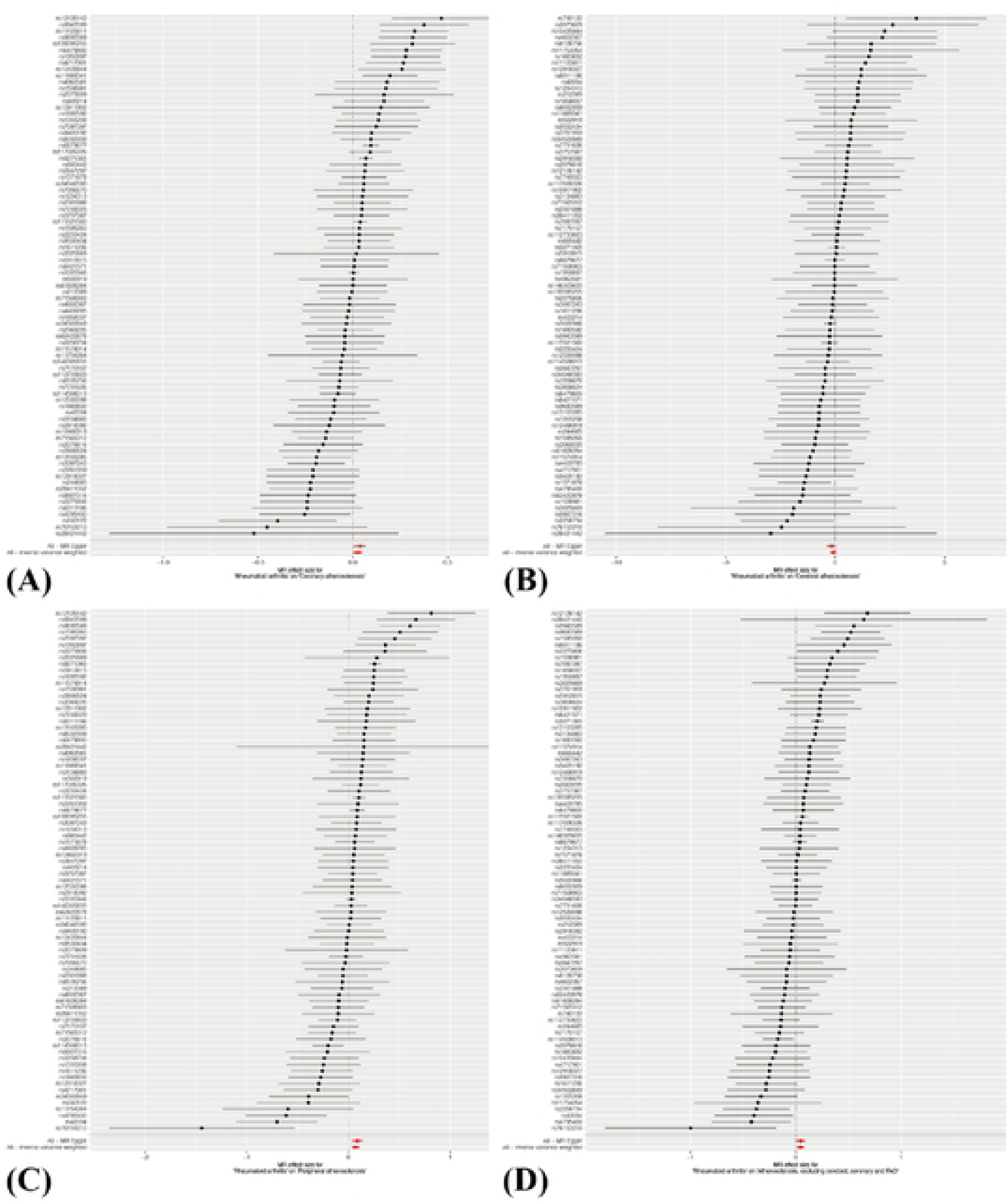
Summary of forest maps of the effects of SNPs associated with RA on AS risk. A: RA on coronary AS. B: RA on cerebral AS. C: RA on peripheral AS. D: RA on ASE. RA, rheumatoid arthritis; AS, atherosclerosis; ASE, atherosclerosis excluding coronary atherosclerosis, cerebral atherosclerosis, and PAD (peripheral arterial disease); SNPs, single nucleotide polymorphisms.

**Supplementary Figure SS.**
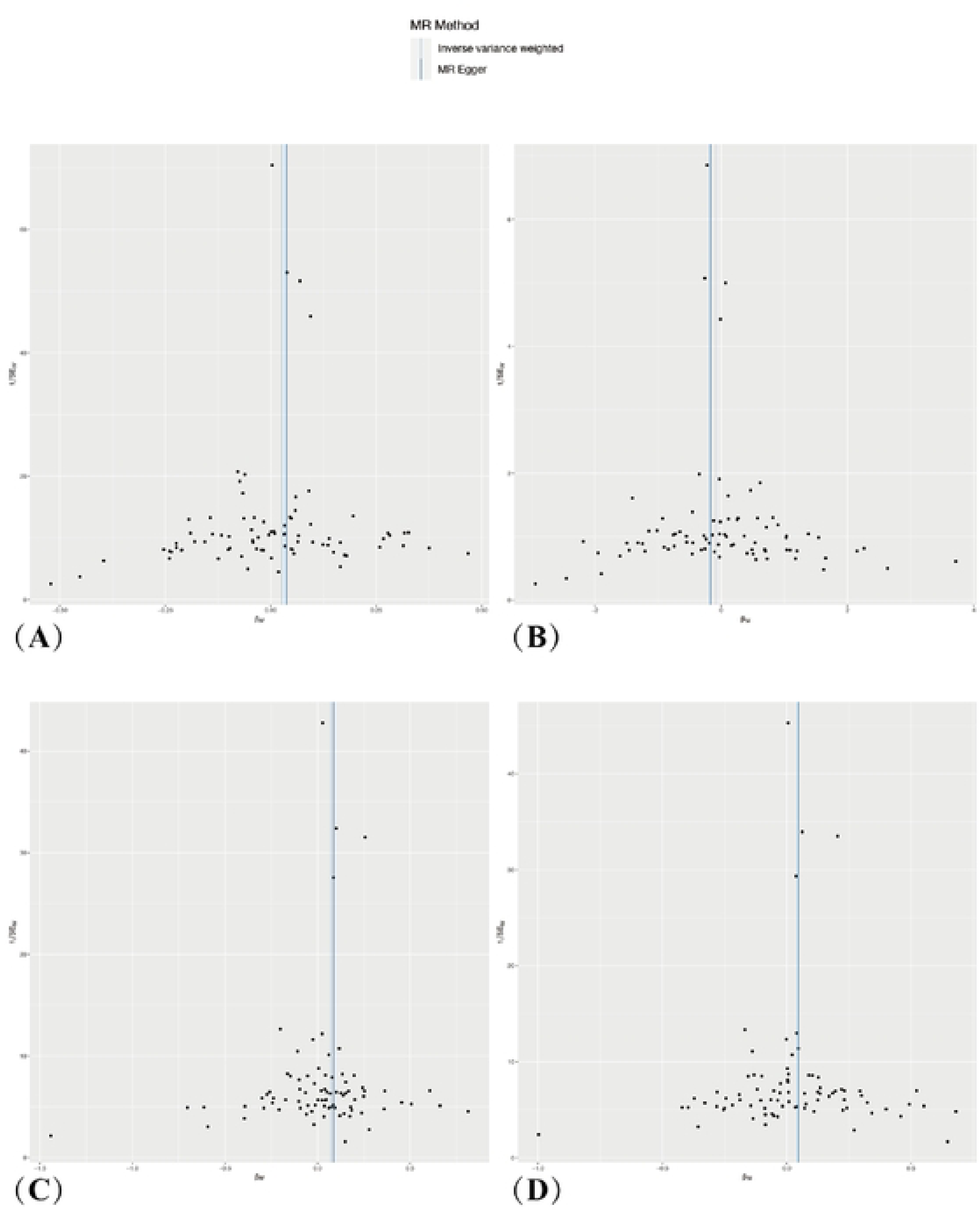
Funnel plot analysis of SNPs associated with RA and AS risk. A: RA on coronary AS B: RA on cerebral AS. C: RA on peripheral AS. D: RA on ASE. RA, rheumatoid arthritis; AS, atherosclerosis; ASE, atberosclerosis excluding coronary atherosclerosis, cerebral atherosclerosis, and PAD (peripheral arterial disease), SNPs, single nucleotide polymorphisms.

**Supplementary Figure S6.**
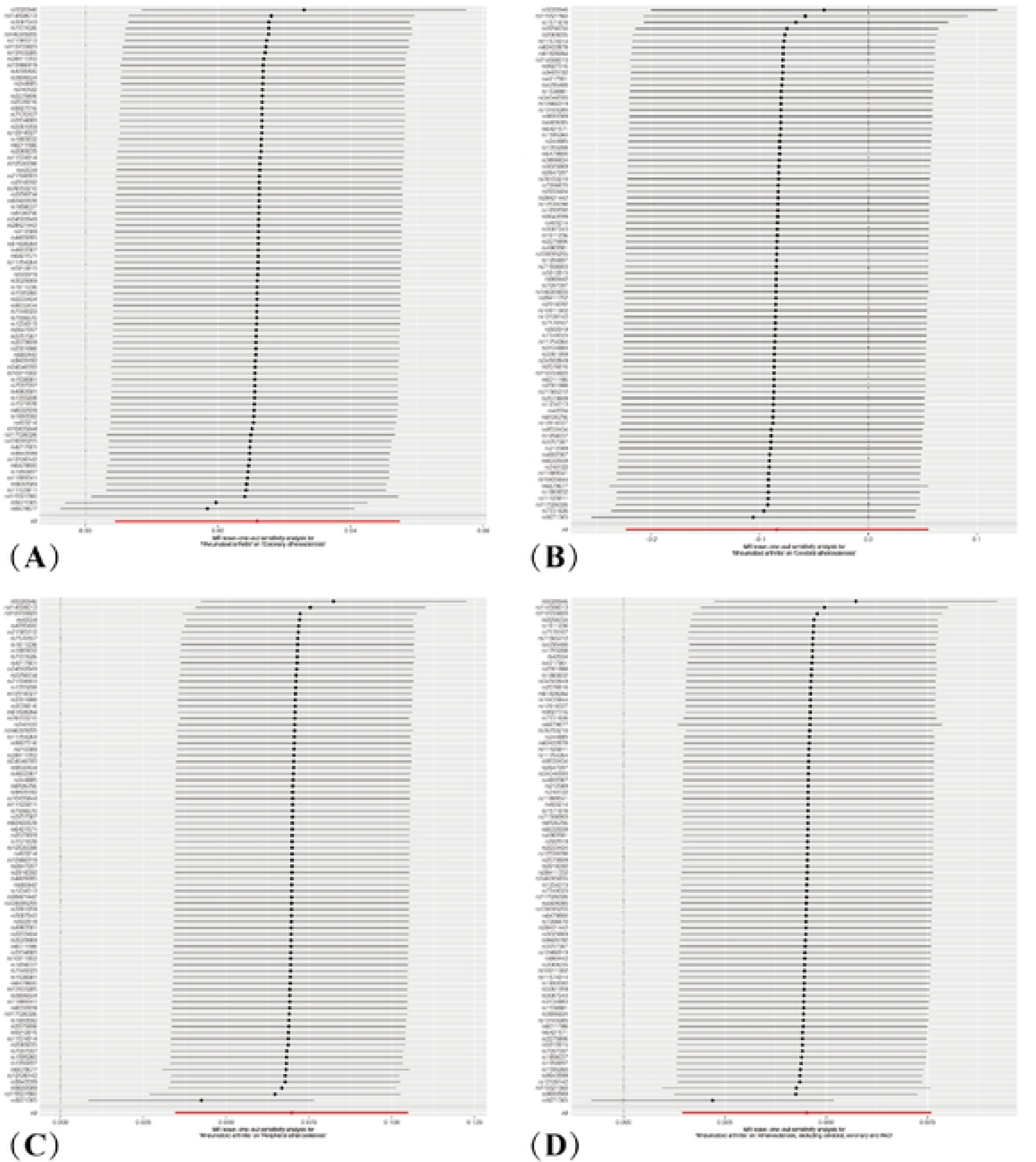
Residual sensitivity analysis of SNPs associated with RA and AS risk. A: RA on coronary AS. B: RA on cerebral AS. C: RA on peripheral AS. D: RA on ASE. RA, rheumatoid arthritis; AS, atherosclerosis; ASE, atherosclerosis excluding coronary atherosclerosis, cerebral atherosclerosis, and PAD (peripheral arterial disease); SNPs, single nucleotide polymorphisms.

**Supplementary Table SI.**
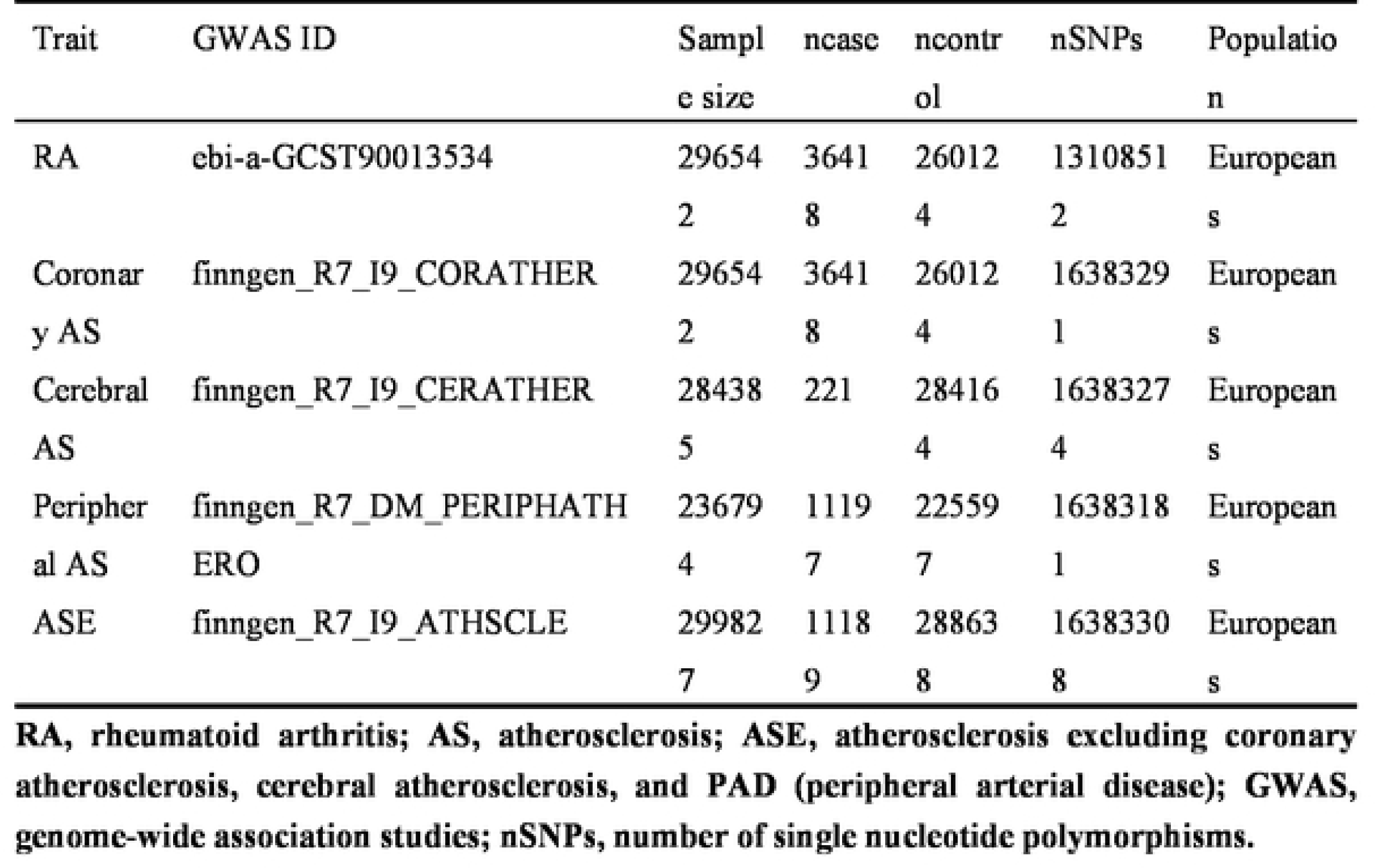
Description of data used for the five phenotypes.

**Supplementary Table S2.** Testing of heterogeneity and pleiotropy of the RA-associated IVs from AS.

| Outcomes | Heterogeneity test |  |  |  |  |  |  |  |  |
| --- | --- | --- | --- | --- | --- | --- | --- | --- | --- |
|  | Inverse-variance weighted |  |  | MR-egger |  |  | MR-egger |  |  |
|  | Q | Q_df | P | Q | Q_df | P | intercept | SE | P-val |
| Coronary AS | 207 | 81 | <0.0001 | 204 | 80 | <0.0001 | -0.0028 | 0.0029 | 0.3408 |
| Cerebral AS | 66 | 81 | 0.8926 | 64 | 80 | 0.8970 | 0.0201 | 0.0186 | 0.2839 |
| Peripheral AS | 205 | 81 | <0.0001 | 203 | 80 | <0.0001 | -0.0040 | 0.0047 | 0.4021 |
| ASE | 175 | 81 | <0.0001 | 175 | 80 | <0.0001 | -0.0006 | 0.0041 | 0.8846 |

**RA, rheumatoid arthritis; AS, atherosclerosis; ASE, atherosclerosis(excluding coronary atherosclerosis, cerebral atherosclerosis and PAD); p, P value; P-val, P value; IVs, instrumental variables; SE, standard error; Q\_df, Q degree of freedom.**

**Supplementary Table S3.** The results of reverse UVMR analysis are based on IVW, MR-Egger regression and WM methods.

| Exposures | Method | nSNPs | P-value | OR | OR_lci95 | OR_uci95 |
| --- | --- | --- | --- | --- | --- | --- |
| Coronary | IVW | 148 | 0.1180 | 0.9509 | 0.8928 | 1.0129 |
| AS | MR-Egger | 148 | 0.4765 | 0.9215 | 0.7362 | 1.1534 |
|  | WM | 148 | 0.8773 | 0.9937 | 0.9165 | 1.0773 |
| Cerebral | IVW | 4 | 0.6242 | 0.9886 | 0.9440 | 1.0352 |
| AS | MR-Egger | 4 | 0.7469 | 0.9679 | 0.8141 | 1.1507 |
|  | WM | 4 | 0.4272 | 0.9756 | 0.9177 | 1.0370 |
| Peripheral | IVW | 81 | 0.6522 | 0.9812 | 0.9032 | 1.0658 |
| AS | MR-Egger | 81 | 0.6107 | 0.9382 | 0.7346 | 1.1982 |
|  | WM | 81 | 0.7128 | 0.9880 | 0.9264 | 1.0537 |
| ASE | IVW | 79 | 0.5138 | 0.9659 | 0.8704 | 1.0719 |
|  | MR-Egger | 79 | 0.6172 | 0.9340 | 0.7153 | 1.2195 |
|  | WM | 79 | 0.6643 | 0.9840 | 0.9150 | 1.0583 |

AS, atherosclerosis; nSNPs, number of single nucleotide polymorphisms; IVW, inverse variance-weighted; WM, weighted median; OR, odds ratio; OR\_lci95, The lower limit of the 95% confidence interval for the odds ratio; OR\_uci95, The upper limit of the 95% confidence interval for the odds ratio; ASE, atherosclerosis excluding coronary atherosclerosis, cerebral atherosclerosis, and PAD (peripheral arterial disease).

**Supplementary Table S4.**
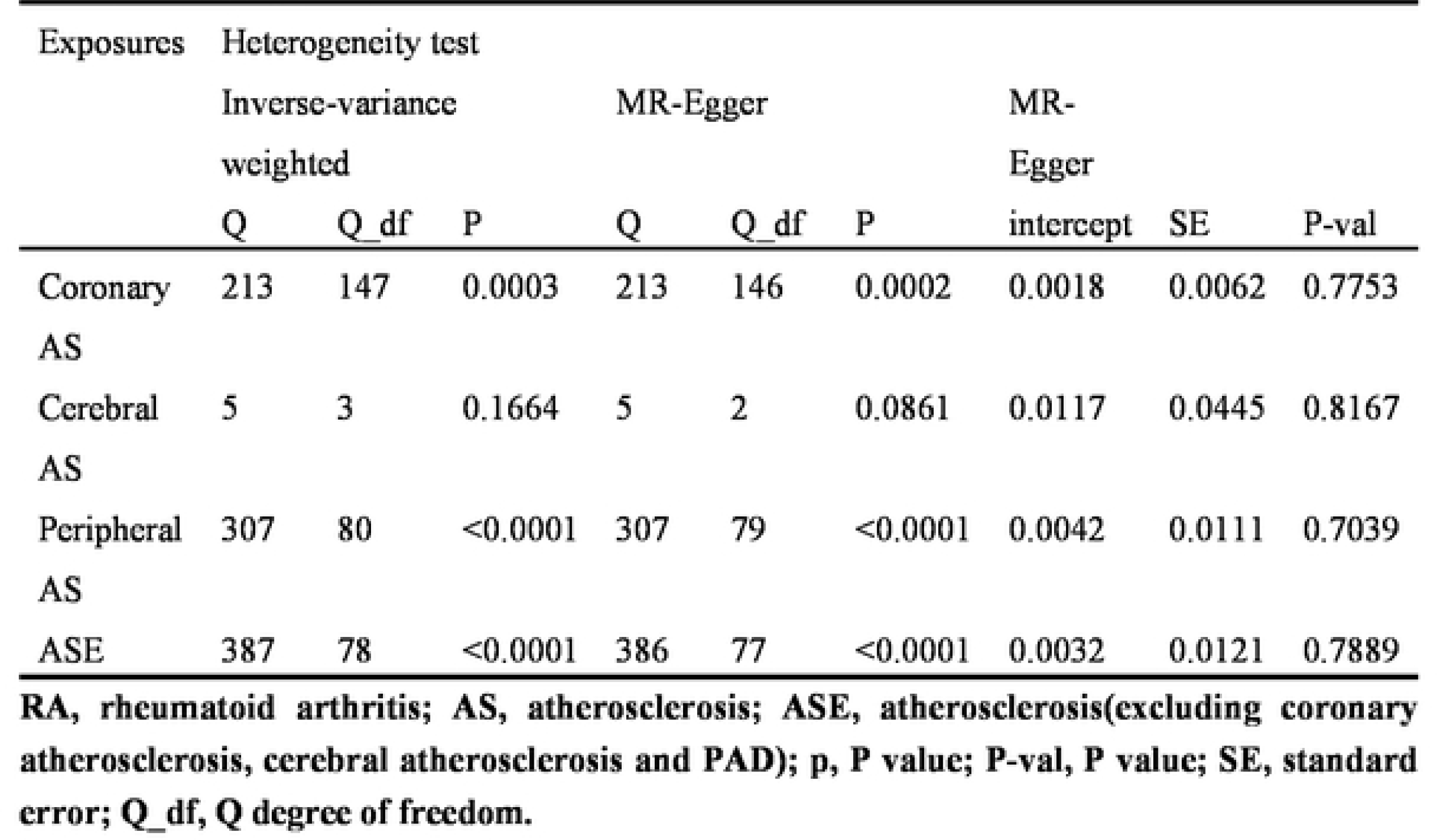
Testing of heterogeneity and pleiotropy of the AS -associated IVs from RA.

